# Dealing with Covid-19 infections in Kolkata, India: A GIS based risk analysis and implications for future scenarios

**DOI:** 10.1101/2020.08.31.20185215

**Authors:** Bibhash Nath, Santanu Majumder, Mohammad Mahmudur Rahman, Jayanta Sen

## Abstract

The Covid-19 pandemic has inherently affected daily lives of people around the world. We, the people, have already started to live in a new-normal way, such as wearing masks, keeping a safe distance from others and maintaining higher level of health & hygiene standards. However, because of this pandemic, the global economy has taken a major blow. Many people have lost their livelihood, and now facing challenges in getting better healthcare facilities and food for survival. Given the serious challenges to the problem, this paper analyzes demographic data to predict vulnerable areas in Kolkata metropolitan city that houses nearly one-third of its population in slums and one-fifth below poverty level, under compromised living conditions. The analysis revealed that the highest risk areas are located in the east and west of the city, the area to a great extent overlapped with wards containing larger share of population below poverty level and are also living in slums. The analysis of publicly accessible Covid-19 case records and containment zones data indicate the hardest hit areas lies in the central Kolkata and several wards along the eastern and northeastern border of the Kolkata Municipal Corporation. The data further revealed that the virus infections have extended to the south Kolkata with increasing number of ‘broad-based’ containment zones with heightened cases. The analysis of demographic characteristics of the hardest hit wards revealed that not a single variables are directly associated with the increase in the number of containments for a particular ward. The ranking of wards based on four intervention criterion have suggested that the ‘lack of social awareness’ along with ‘lack of social distancing’ have dominantly contributed to the increasing number of containments of Covid-19 cases in Kolkata. Determination of optimized ranking and Spearman’s rank correlation coefficient of the wards based on four intervention criterion provided a basis for the policy makers to assess ward-based interventions criterion to control further spread of the disease and/or prevent second wave of infections. Given that the effective antiviral drugs is far away from common publics reach, the application of our study approach would benefit saving lives of many vulnerable populations.

## 1. Introduction

The entire world is currently grappling with deadly infections from a novel coronavirus disease 2019 (Covid-19) with as many as 24 million people globally have been infected and a total global deaths of 0.81 million people (https://coronavirus.jhu.edu/). The World Health Organization (WHO) have declared this catastrophe as a pandemic in March 2020. The disease has a flue like symptoms and affects each individual differently.

Covid-19 is a new phenomenon with a variety of risk factors and many remain unknown. Many social determinants of health (SDoH), such as housing, physical environment, food and poverty, can have a detrimental effect on Covid-19 infections and mortality (Abrams and Szefler, 2020). Several potential risk factors have been identified including: age, race/ethnicity, preexisting medical conditions, poverty, crowding, hygiene, and certain occupations (https://www.cdc.gov/). In addition to that poor living conditions, lack of clean drinking water and sanitation could increase the risk to the problem. Lack of access to quality healthcare would add further to contraction and transmission of the disease. Homeless people living in shelters or in streets can have a much higher risk of contracting the virus due to their inability to maintain social distancing and possibility of not having basic hygiene supplies (Baggett et al., 2020; Tsai and Wilson, 2020). Therefore, it is an urgent need for the scientific community to carry out risk assessment based on demographics and help policy makers to make an appropriate prevention and/or intervention strategies to allocate resources to the most vulnerable areas, i.e., the areas with high risk and high needs. Such an approach would help control the transmission of the disease, and minimizes the impact on our already stressed existing healthcare infrastructures.

Covid-19 infections have shown no barriers in terms of geography, race, ethnicity and wealth. Among the hard hit countries USA, Brazil, India and Russia themselves share the burden of over 50% of all positive cases. Due to this high volume of infected people, the health infrastructure in these countries is at its breaking point. Scientists around the world are working tirelessly to find a cure for this disease and it might take a considerable time before the general public would get a chance for vaccination. Therefore a focused planning is absolutely essential to protect people from contracting Covid-19 in the future.

India is one of the poorest countries in the world with a total population of 1.35 billion. So far 3.1 million people have been infected and with over 50,000 new cases adding daily. India ranks low on social mobility index (76^th^ out of 82 countries; WEF, 2020) with about 22% population live below poverty level (BPL) and a literacy rate of 74% (https://censusindia.gov.in/). Large section of the workforce are in the unorganized sectors and many are on daily-wage and migrant labors. The implementation of various prevention program and lockdown measures that have initially helped India to slow the spread of virus. However, such tough measures have impacted the economy badly in almost every sectors and especially the unorganized sectors. Therefore, prevention and/or intervention strategies should be adopted by taking into consideration the need and necessity of underprivileged population. For example, the U.S and other countries, the government have taken various humanitarian steps and taken care of the vulnerable population by providing foods, shelters, and stimulus package during this critical time of economic shutdown (https://www.usa.gov/coronavirus).

Kolkata is one of the hard hit cities in India with a total Covid-19 infections of over 37,000 and total deaths of 1,200 (https://www.covid19india.org/). The situation is at the critical level in most part of the city sprawling north and central Kolkata, and more recently many people are getting infected in south Kolkata. Kolkata Municipal Corporation (KMC), with 141 wards, has a total of 4.5 million people and roughly 30% of them are living in slums where living standards are highly poor (Ghosh, 2013). According to the report of Health & Family Welfare Department of Government of West Bengal, some geographical areas in the State are reporting higher number of Covid-19 cases and most of these cases are actually from few pockets/settlements and/or families. Thus, identification of those pockets or settlements and contained them was an important first step of gaining an upper hand from the virus transmission. The Government of West Bengal has followed a containment strategy principles to break the chain of transmission and preventing the virus from spreading (https://wb.gov.in/COVID-19.aspx). These containment zones are of three types: broad-based which are typically a locality where the number of infections are many, isolation zones/units where the number of infections are one to many, and standalone houses/premises where the number of infections are one to a few. The Government of West Bengal has made this detailed list public until the end of June 2020 (https://wb.gov.in/COVID-19.aspx). However, from the beginning of July 2020, only broad-based containment zones are being reported. The government has not made public the ward wise list of total infections within the KMC. We therefore accessed those containment zone listings to estimate approximate number of positive Covid-19 cases in various KMC wards and also get an understanding of the distribution of containment zones.

In this paper, we analyzed different risk factors associated with Covid-19 infections based on demographics and other variables to identify the areas of high risk for contracting Covid-19. The risk map would be used to make preventive action plan for a sub-city (ward) level in the event of a fresh outbreak. In addition to that we analyzed containment zones listings data to identify the hard hit locations within the KMC during the current wave. The objectives are: 1) To identify locations of the highest risk of contracting Covid-19, and 2) To identify locations of the hardest hit areas of Covid-19 infections within the KMC. Given the uncertain nature of the problem, the identification of both high risk and hard hit areas within the KMC would benefit the policy makers to enable them to address and/or formulate various prevention and/or intervention program based on risk criterion and methodologies that have been summarized in this paper.

## 2. Data collection and methodology

### 2.1 Data used

The data were collected from the Population Enumeration Primary Census Abstract and Houselisting and Housing Census Primary Census Abstract, 2011 of the Government of India (https://censusindia.gov.in/). The ward wise data for Kolkata Municipality Corporation (KMC) were tabulated for total households, total population, total uneducated population, total marginal workers, total number of rooms in the household, total number of persons in the household, and the availability of wastewater drain, water sources and latrine in the household. We have computed ward wise population below poverty level and slums from Basu (2015) and Das Gupta, 2009). The data on homeless people sleeping in the street were calculated from Sabuj Sangha (2014). We then computed total population per households in each wards by dividing total population with total households for those wards. Additionally, we have computed population density in each wards. As a secondary information for discussion purpose we have used average arsenic (As) concentration in untreated groundwater used for drinking and cancer risk within KMC wards from Chakraborti et al. (2017). Tuberculosis (TB) burden data were adapted from Dey et al. (2019). The description of demographic and other variables used in the risk analysis are detailed in Table 1.

**Table 1.** Descriptions of demographic and other variables used in the analysis and discussion.

| **Risk criterion** | **Variables** | **Descriptions (ward-wise)** |
| --- | --- | --- |
| Lack of social distancing<br>(Transmission risk) | Population density | People per Km<sup>2</sup> |
|  | Large family (9+ people) residential households | Percentage of household to total households |
|  | No exclusive room | Percentage of household to total households |
|  | Drinking water sources – far away from home | Percentage of household to total households |
| Lack of Covid-19 awareness | Uneducated population | Percentage of uneducated population |
|  | Slum population | Percentage of population living in slums |
| Susceptible population and/or vulnerable groups | Households below poverty level | Percentage of household to total households |
|  | Marginal workers | Percentage of marginal workers |
|  | Homeless sleeping in streets<sup>1</sup> | Percentage homeless people |
| Poor health & hygiene condition | Untreated water sources | Percentage of household to total households |
|  | Community latrine facility | Percentage of household to total households |
|  | Open/No wastewater drains | Percentage of household to total households |
| Other variables | Arsenic in drinking water sources<sup>1</sup> | Average concentration |
|  | Cancer risk<sup>1</sup> | Calculated from average As concentrations and consumption rate |
|  | Tuberculosis (TB) burden<sup>2</sup> | High or Low |

*Data source:* Census 2011 of India. <sup>1</sup>Homeless people sleeping in streets are from Sabuj Sangha (2014),
<sup>2</sup>Arsenic in untreated groundwater used for drinking and cancer risk from Chakraborti et al. (2017), and
<sup>3</sup>Tuberculosis (TB) burden is from Dey et al. (2019).

### 2.2 Cosine similarity index

We used cosine similarity index for Covid-19 risk analysis using demographic variables. In this method, the relevant input variables were standardized first involving a z-transformation. This means that all the input values (X) were subtracted from the mean (X) for all values and then divided by the standard deviation (σ), to keep all the attributes on the same scale. The method follows cosine similarity mathematics and compares two vectors, i.e., vectors of standardized data for each candidate features (i.e., a municipality ward) and the vectors of standardized attributes for the target feature (a worst-case scenario based on potential risk factors for Covid-19 contraction) to be matched. The cosine similarity of two vectors, A and B, is then computed using the following equation:

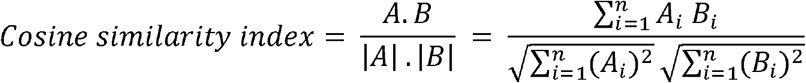

In Cosine similarity index, the relationships among the attributes were considered instead of the attributes magnitudes. The cosine similarity index ranges between 1.0 (perfect similarity) and −1.0 (perfect dissimilarity). The analysis has been performed using ‘Similarity Search’ tool in ArcGIS Pro 2.4 (ESRI). The similarity search tool find other features similar to a worst case scenario in terms of input variables. Initially, a hypothetical ‘target feature’ was created by giving attributes of all the worst values of the variables used in the risk analysis. The variables include: population density, percentage of households with no-exclusive bedrooms, percentage of households with more than 9 members, percentage of households depending on community water source, percentage of uneducated population, percentage of slum population, percentage of marginal workers, percentage of population below poverty level, percentage of houseless population sleeping in the street, percentage of households drinking water from untreated source, percentage of households use community latrine, and percentage of households with no wastewater drains.

Similarity search tool rank all the wards based on ‘attribute similarity’ in comparison with the worst-case scenario, i.e., a ‘target feature’. The ward with the lowest ranking indicates the highest risk, while the ward with the highest ranking indicates the lowest risk for contracting Covid-19. These rankings have been later reversed to align the rank order by giving largest rank to highest risk ward and smallest rank to lowest risk ward.

### 2.3 Risk indices and intervention criterion

We have calculated several risk indices and intervention criterion by normalizing the input data that allowed to assess inter-ward variability in specific Covid-19 risk criterion. These area: social distancing index, social awareness index, susceptibility index, and health & hygiene index. Larger index values indicate higher risk, e.g., a ward with largest social distancing index would ranked the highest, suggesting that the social distancing is lacking, i.e., all indices are positively associated with the Covid-19 virus.

Social distancing and/or physical isolation is one of the most important factor that controls the transmission of the virus (Kaur et al., 2020). The following variables have been used in the calculation of social distancing indices, these are – population density (i.e., the number of people per km^2^), household size (i.e., the percentage of households with 9+ people), availability of exclusive bed room in the household (in percentage), and the percentage of household access community drinking water sources. We considered that 9+ people in a household would make it difficult to maintain social distancing and/or physical isolation in case one of the family member gets infected. Similarly non-availability of exclusive bedroom would make it difficult to maintain physical isolation. This is also true for a household member that had to collect drinking water from community wells or timed tap water sources. The chances of social distancing and/or physical isolation would be compromised during drinking water collection.

For social awareness index, we have used demographic variables such as percentage of population living in slums and percentage of uneducated population. We believe that the raising of community awareness would benefit the purpose of social distancing because if someone is not aware of the virus and how the transmission/contraction can occur would risks self and others because the transmission of Covid-19 virus can occur by indirect contacts with surfaces or objects used by an infected person (Jayaweera et al., 2020). Therefore community educating would essentially help keeping the infection under control.

For susceptibility index, we have used percentage of BPL households, percentage of homeless people sleeping in the streets and percentage of marginal workers. This index would help addressing the needs of most vulnerable population in the community. Because during a pandemic and economic lockdown, most of these vulnerable group of people would be out of jobs. Therefore, it would be essential to help these people so that they have the food for survival and otherwise it would be difficult for them to participate in the broader cause of containing the virus. If we do not consider this index than there would be a possibility of a major outbreak of virus infections in those areas.

Lastly, we have calculated health & hygiene index to address vulnerable areas where people have the possibly of having weak immune system. In this index, we have considered percentage of households drinking water from untreated sources (typically contain pathogens, toxic trace elements such as As, Fe and Mn, and chances of waterborne viruses; Chakraborti et al., 2017), percentage of households with no latrines within the premises (community latrine usually have poor hygiene condition), and percentage of households have no wastewater drains.

The ward wise data of each variables were first normalized (*X_normalized_*) to create an index for each variables, followed by ranking of the wards by giving largest rank for highest index values. Finally a composite index (*X_CI_*) was calculated based on rank normalization.

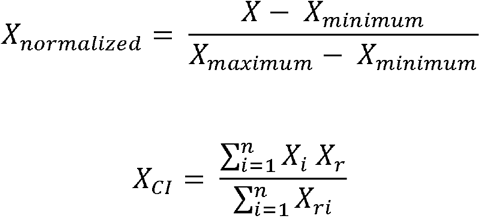

The composite indices (or intervention criterion) for four criterion were again ranked by giving largest rank for highest index values for different wards.

We then created a ward wise predominance map based on the ranks of four criterion (i.e., social distancing, susceptibility, social awareness and health & hygiene) to show which of the four criterion has the highest intra-ward rank. For example, if a ward has the social distancing rank of 136, susceptibility rank of 123, social awareness rank of 105, and health & hygiene rank of 70 than we assigned social distancing as the predominant category based on the rank value. We also calculated the strength of this predominant category using rank difference (i.e., the difference between 1^st^ and 2^nd^ category rank value) and percentage total of rank value. Higher the percentage total rank value and/or the rank difference the more predominant the category would be. This map was created to determine the intervention category that would be better suited for different KMC wards during the current cycle of Covid-19 infections.

Finally, Spearman’s rank correlation coefficient was used to derive ward wise ‘optimum prevention rank’ based on social distancing, susceptibility, and social awareness, and health & hygiene conditions. We used different combination of weight factors given to each of the risk criterion for the calculation of ‘optimum prevention ranking’. These optimum wards rankings allowed policy makers to select a set of largest ranked wards for priority prevention. The Spearman’s correlation (*ρ*) was calculated using the following equation:

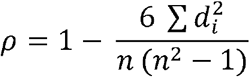

Where, *d_i_* = difference in paired ranks and *n* = number of cases.

## 3. Results and discussion

### 3.1 Covid-19 risk mapping

The cosine similarity index (CSI) calculated based on attribute profile were used to rank all KMC wards. The highest risk wards (i.e., a similarity rank between 1 and 30) are mostly clustered to the east and west of the KMC area (Fig. 1). However, several high risk wards (i.e., a similarity rank between 31 and 60) are scattered throughout the KMC area covering both north and south part. The spatial pattern of risk map suggests a widespread nature of Covid-19 risk to population residing within the KMC. This risk map forms a basis for further analysis of demographic variables to help prioritize prevention strategies and providing a suitable planning scenarios to achieve the maximum benefits. This is highly likely that different demographic variables (i.e., various risk factors) will act differently in different regions of the KMC area.

**Figure 1.**
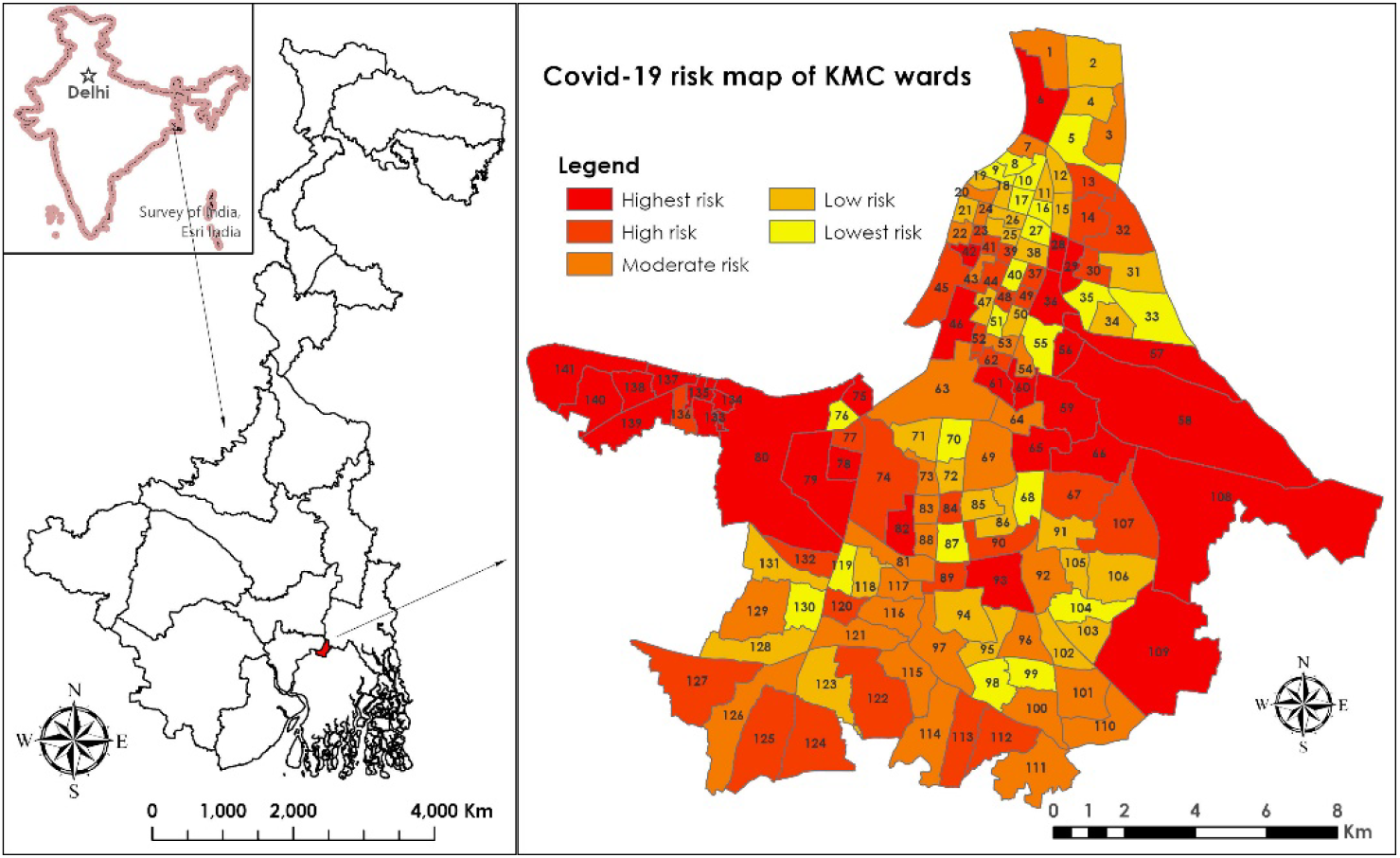
Map of the study location in India, showing Covid-19 risk based on multiple demographic variables. The risk areas were identified on the basis of a ‘similarity search’ in ArcGIS Pro 2.5 (ESRI) using attribute profile and calculation of Cosine similarity index (CSI) for 141 wards of Kolkata Municipal Corporation.

The analysis of individual variables showed that the population density of KMC wards is the highest in north Kolkata and some pockets in central and west Kolkata (Fig. 2a). Overall, the population density in south Kolkata is low (<25,000 people/Km^2^). As such population density is not directly linked to Covid-19 infections unless social distancing guidelines are followed, according to the study of over 900 metropolitan counties in the US (Hamidi et al., 2020). This is also true for New York City, where majority of the infected population are from the low income zip codes and living in the crowded households (Nath, 2020). Therefore, we believe that the population density, if not alone, together with other variables such as large family household, lack of social awareness, and reluctant to follow social distancing guidelines should play a major role in the virus transmission. The large family household are mostly observed in the west Kolkata near Garden Reach and Metiabruz region and parts of north Kolkata, while in south Kolkata <3% households have more than nine people in comparison to west and parts of north Kolkata where >6% households share their space with more than 9 people (Fig. 2b). Therefore, the combination of these two variables should have contributed to the higher risk of Covid-19 infections in the west and north Kolkata. Because, maintaining a safe physical distance is a top criterion in these areas because if someone from a crowded household gets infected there would be difficultly for the other members to maintain a safe distance from the infected person or if there is an asymptotic persons in the household the chances of risk would increase significantly in comparison to a small family households. Higher percentage of households below poverty level (BPL) households can be found in several pockets in the east and west of Kolkata (Fig. 2c). These areas also houses larger percentage of population in slums (Fig. 2d). In addition to that higher percentage of uneducated population (>25% of the total population) are closely associated with the people living in slums and below poverty level (Fig. S11a). However, the higher percentage of marginal workers (>6% of total population) are found in 24 wards throughout the KMC (Fig. S1b). In several wards, people use community latrine which could be a risk factor on poor hygiene condition (Fig. S1c). Thus, it would be essential to make a consideration in association with BPL population, marginal workers and slums because these socially excluded population lacks space and physical environment as well as lack access to clean drinking water, sanitation and hygiene condition (Mayne, 2017). In south Kolkata, people are at risk of drinking water from untreated sources, while in north Kolkata >90% households have access to clean water. According to a study on groundwater quality, high concentrations of As above WHO guidelines of 10 μg/l are largely found in the groundwater sources from southern KMC (Fig. S2a; Chakraborti et al., 2017). As such drinking of As contaminated water for a long time could cause carcinogenicity and thus effects immune system. Additionally, Covid-19 virus potentially sourced from untreated wastewater (Quilliam et al., 2020). As such larger percentage of households in south Kolkata either do not have wastewater drains or having open drains. Therefore, the hygiene conditions could be under compromised conditions in those areas. We speculate that the case-morbidity rates is much higher in south Kolkata than in other areas of KMC. Additionally, large parts of east and south Kolkata together several pockets in north and west Kolkata coincides with high Tuberculosis (TB) burden (Fig. S2b; Dey at al., 2019), suggesting a lack of health and hygiene conditions in those areas. Studies showed that the Tuberculosis (TB) is associated with a 2.1-fold increased risk of a severe COVID-19 disease (https://www.mohfw.gov.in/), and thereby India recommended bi-directional TB-COVID screening. According to World Health Organization (WHO) report, Covid-19 is expected to affect patients with tuberculosis (TB). However, the responses of Covid-19 and TB complement each other (Dara et al., 2020) which could be a positive sign for Kolkata slums where the population should already have the basic education on TB etiquettes and thus the similar etiquettes could be applied to prevent Covid-19 risk.

A preventive plan could be deduced based on the analysis of risk factors and demographic variables, while selection of specific risk factors would require prioritization such as raising awareness among uneducated population and teaching social distancing norms to those living in dense areas. A clear prevention efforts would stop Covid-19 infections from occurring.

**Figure 2.**
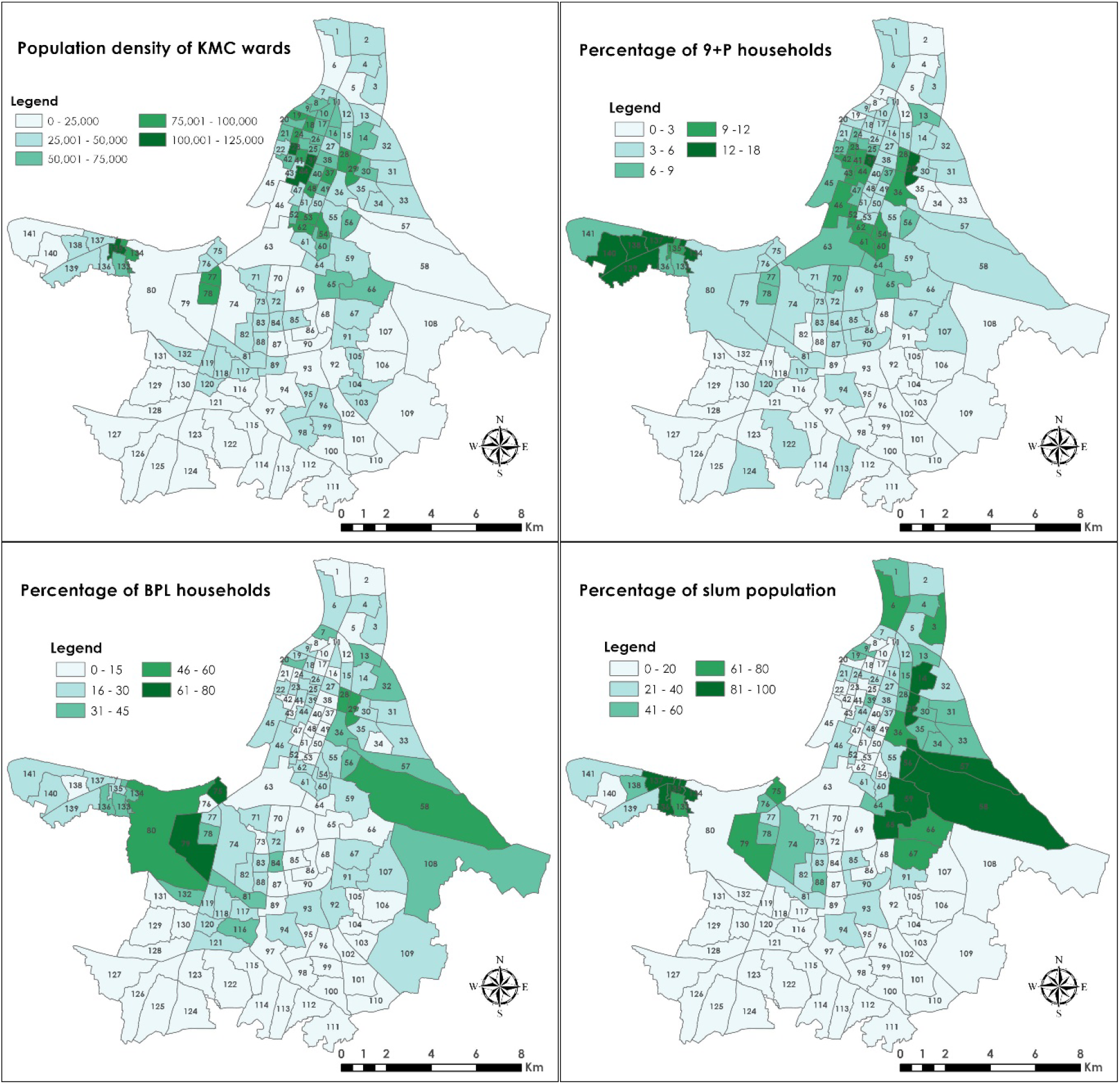
Ward wise distribution of demographic characteristics: A) population density (/Km^2^), B) percentage of 9+ person households, C) percentage of below poverty level (BPL) households, and D) percentage of people living in slums in Kolkata Municipal Corporation.

### 3.2 Identification of hard hit area*s*

The KMC hast not made public the ward wise data on total positive Covid-19 cases till date. Therefore, in order to understand the status of Covid-19 infections and the role of demographic variables (risk factors) in different KMC wards, we analyzed record of publicly accessible containment zones and/or isolation units (https://wb.gov.in/COVID-19.aspx). These containment zones and/or isolation units were established when one or more positive Covid-19 cases were reported from that zones/units and kept that zones/units in isolation for a continuous 14 days of no case reports (https://wb.gov.in/). This has been done to control the transmission of the virus.

We analyzed a total of 3,288 Covid-19 case records from 17 March to 30 June 2020. These data have been accessed from leading newspaper from Kolkata and Twitter/Facebook account of Kolkata Police and similar other governmental sources (https://twitter.com/KolkataPolice/status/1259803430677123072/photo/1). From these records we estimated the total number of infected people in different wards of KMC, by assigning 1 positive case for each record (the most conservative approach). We could estimate roughly about 60% of the actual positive cases within KMC until 30 June 2020. This indicate that some of the case records could be missing from our database or there could be multiple infected persons for one records. According to our estimate, there are at least 43 wards within KMC has at least ≥25 positive cases (Fig. 3a). These wards are distributed in the central and eastern part of the city. The number of wards with ≥25 positive cases would likely increase once all positive cases are accounted for different wards. The analysis of the data further revealed that the containment zones/isolation units have increased over time, from 227 zones in April 27 to 724 zones in June 8. In April 27, more than 4 containment zones/isolation units are found in 12 wards located in central and north Kolkata, while that number grew by 21 wards in May 11 and spread towards east and west of the city. In June 8, 61 wards had more than 4 containment zones/isolation units. This suggests the spatial extent of virus transmission even with a strict containment measures and isolation policy of the positive cases together with nation-wide ‘complete’ lockdown that was in force until 31 May 2020. According to our analysis, the intensity of Covid-19 case records (i.e., number of cases per 1,000 people) is significantly higher in several wards in central Kolkata and a few wards in northeast Kolkata. This suggests a hotspot of heightened cases in the central Kolkata region (Fig. 3b).

**Figure 3.**
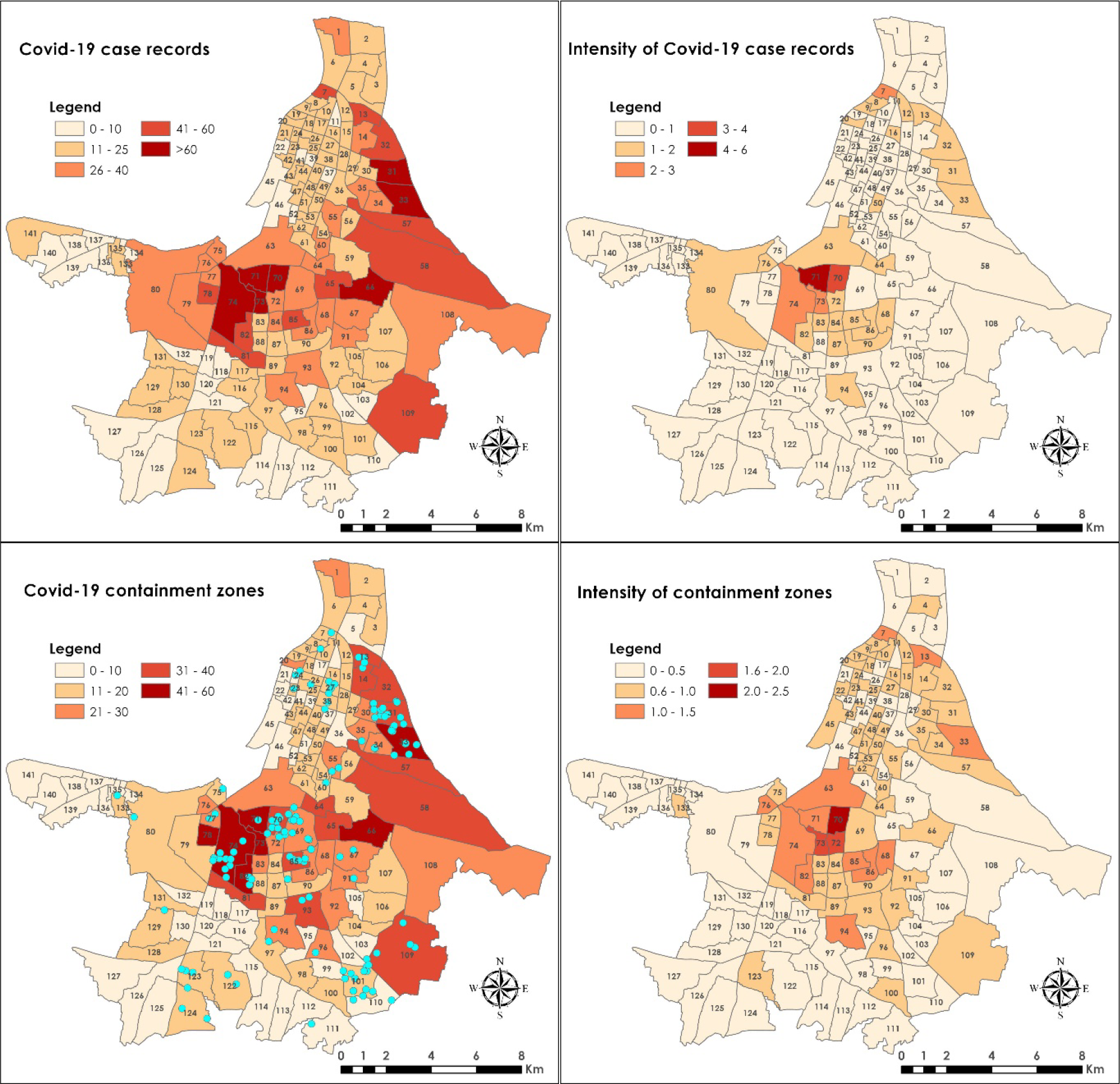
A) Ward wise distribution of total Covid-19 case records until June 30, within the Kolkata Municipal Corporation (KMC). B) Intensity of Covid-19 case records (per 1,000 population) in different wards. C) Ward wise distribution of total Covid-19 containment zones on the basis of over 2,400 containment zone listings until June 30, 2020. D) Intensity of containment zones (per 1,000 population) illustrate the per capita distribution that helps identifying the worst hit locations. Broad-based containment zones since July 1 (total 135) is shown as a point symbol in cyan color and are not included in B) and D), because these containment zones were of redefined nature.

We then removed identical records from the database of 3,288 cases, because one containment zones and/or isolation units is found contain multiple records, to compile information on the localities that have been contained over the three months periods. The summarized data revealed a total 2,413 locations that have contained between 27 April and 30 June 2020. However, since the beginning of July, Government of West Bengal have contained only the locations with heightened cases and redefined containment zones by adding a buffer zone. These zones are kept under total lockdown for 14 days with no cases. Thus, these locations are not reflective of the location of all cases. However, previously they had three categories of such zones, such as affected zones (listed as containment zones or isolation units), buffer zones, and clean zones. Our analysis based on containment zone record identifies 9 wards that have been contained in ≥40 locations over 3 month’s periods. These are ward no. 70, 71, 73, 74, 78 and 82 covering parts of Bhowanipore, Alipore, Ekbalpore, Mominpore, Chetla and Kalighat in central Kolkata, ward no. 66 covering parts of Tangra and Topsia in central Kolkata. In addition, ward no. 33 in north Kolkata covering parts of Phoolbagan and Beliaghata region, and ward no. 109 in south Kolkata covering parts of Ajoy Nagar, Hiland Park and Chak Garia. However, July and August ‘broad-based’ containment zones were shown to scatter in three main locations, central, northeast and southeast Kolkata. These zones largely overlapped with areas that have contained earlier suggesting the difficult nature of the problem where human participation is the key to fight against Covid-19 (Fig. 3c). The data further revealed that the most contained areas within the KMC also have the most case records. Both the ward wise number of records and the number of containment zones are positively correlated with a R^2^ value of 0.92 (after removing an outlier hotspot) (Fig. S3). Times of India newspaper reported on May 1, 2020 that Central Kolkata and pockets of south-east Kolkata are among the localities to bother for KMC officials and Kolkata Police (https://timesofindia.indiatimes.com/city/kolkata/city-has-264-high-risk-spots-new-pockets-in-central-south-east-localities-worry-cops/articleshow/75479992.cms). However, the highest intensity containment zones (i.e., >1 containment zones per 1,000 population) are found in ward no. 70, 71, 72 and 73 and their adjacent wards of central Kolkata, while other regions have significantly lower intensity of containment zones (Fig. 3d). In total, 15 wards of KMC have the intensity >1 containment zones per 1,000 population and thus could be considered as a major Covid-19 hot spot in KMC.

### 3.3 Community characteristics of hard hit areas

It is important to analyze community characteristics of hard hit wards before making a plan for intervention so that resources will be ready in the event of a second wave of virus infections and/or controlling the further transmission of the virus. The analysis of demographic characteristics will help understanding the needs of people in different hard hit localities and addressing the specific needs of population, such as food and shelters for the poor and homeless, availability of health resources, social distancing guidelines, distribution of masks and community awareness program.

Analysis of the community characteristics showed that not a single demographic variables are showing strong positive correlation with the number of containment zones (Fig. S4), suggesting that a combination of demographic factors could be responsible for the spread of virus infections. For examples, we find five wards with low population density (<50,000 people per Km^2^) but the number of containment zone is much higher (>40). While, we find sixteen wards with high population density (>75,000 people per Km^2^) but the total containment zones are <20. This suggests that population density is not a dominant factor for virus transmission, also suggested by others (e.g., Hamidi et al., 2020). This is also true for households with more than 9 people. In addition to that, we observed an inverse relationship between the percentage of HHs using untreated water and containment zones, and percentage of HHs with no wastewater drains and containment zones. We found that the relationship between total marginal workers and containment zone is somewhat positive. Similar relationship pattern is also observed for the percentage of BPL HHs, total uneducated population, and total population in slums with total containment zones.

We then chose 15 hard hit wards where the intensity of containment zones were >1 per 1,000 population and analyzed demographic characteristics, so that inter-ward comparison can made to understand the reason behind the observed hotspots (Fig. 4). The analysis revealed that in three wards (ward no. 7, 63 and 72) >8% households have the non-availability of exclusive bedrooms, while in four wards (ward no. 13, 63, 70 and 71) >6% households are crowded with 9+ persons (Fig. 4a). In addition to that, in six wards (ward no. 7, 13, 73, 76, 82 and 85) population density is >40,000 people per km^2^ (figure not shown). Among other variables, 6 wards (ward no. 7, 13, 33, 74, 76 and 85) have higher share of population living in slums (>30% people). In 3 wards (ward no. 7, 13 and 82) >25% households are below poverty level. In addition to that, in five wards (ward no. 7, 68, 82, 85, 86 and 94) >7% households have only access to community latrines, while 1 ward (ward no. 13) have >5% households have only access to untreated water sources.

**Figure 4.**
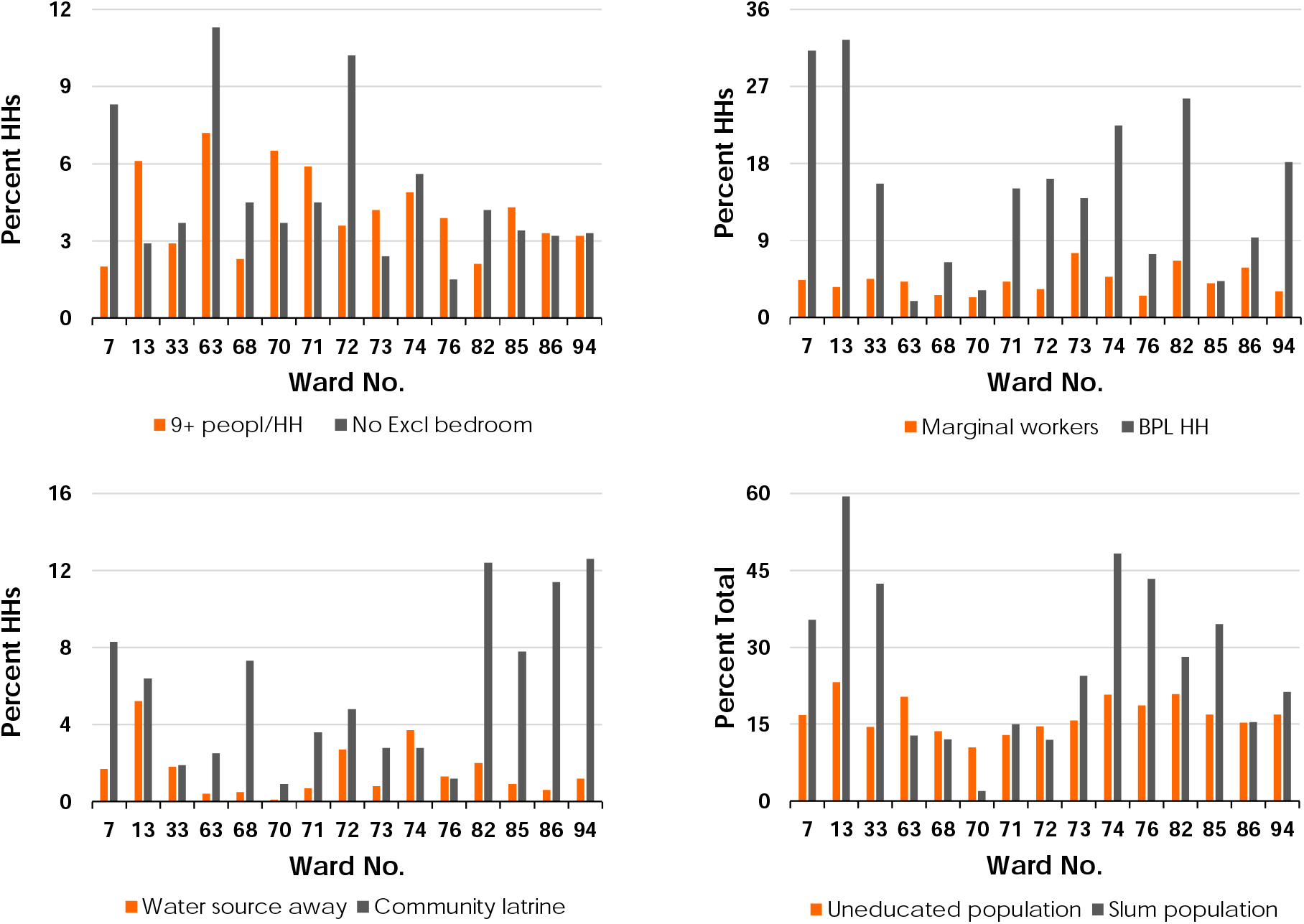
Demographic characteristics of fifteen hard hit wards (>1 containment zone per 1,000 population) of Kolkata Municipal Corporation.

The above analysis suggests that the community characteristics are different in different hard hit wards. Therefore, it is important to formulate multiple planning scenarios associated with the community characteristics to achieve the best results.

### 3.4 Rank weighted rankings of wards based on intervention criterion

Based on the analysis of demographic characteristics for the entire KMC and hard hit wards four criteria based planning objectives were made. These are lack of social distancing, lack of community awareness, susceptibility and poor health & hygiene conditions. Based on these criterion, all the wards were ranked using respective index values, highest index value ranked largest and vice versa.

According to social distancing rank map (i.e., the largest rank indicates the highest priority to follow social distancing guidelines), most of the largest ranked wards are located as a clusters of wards in north Kolkata, several wards in the west near Garden Reach and Metiabruz areas, and a few wards in eastern Kolkata. These are the areas having highest population density, higher percentage of large family (9+ people) households, and water sources far way (Figs. 5a and S1). Spearman’s rank correlation coefficient has been applied to determine which variable has a more important role on the rank weighted social distancing index rank. The *rR* values indicate that the population density (*r_R_* = +0.82) and crowded households (*r_R_* = +0.5) have played an important role on the ‘optimum rank’ than non-availability of exclusive rooms and location of water source.

**Figure 5.**
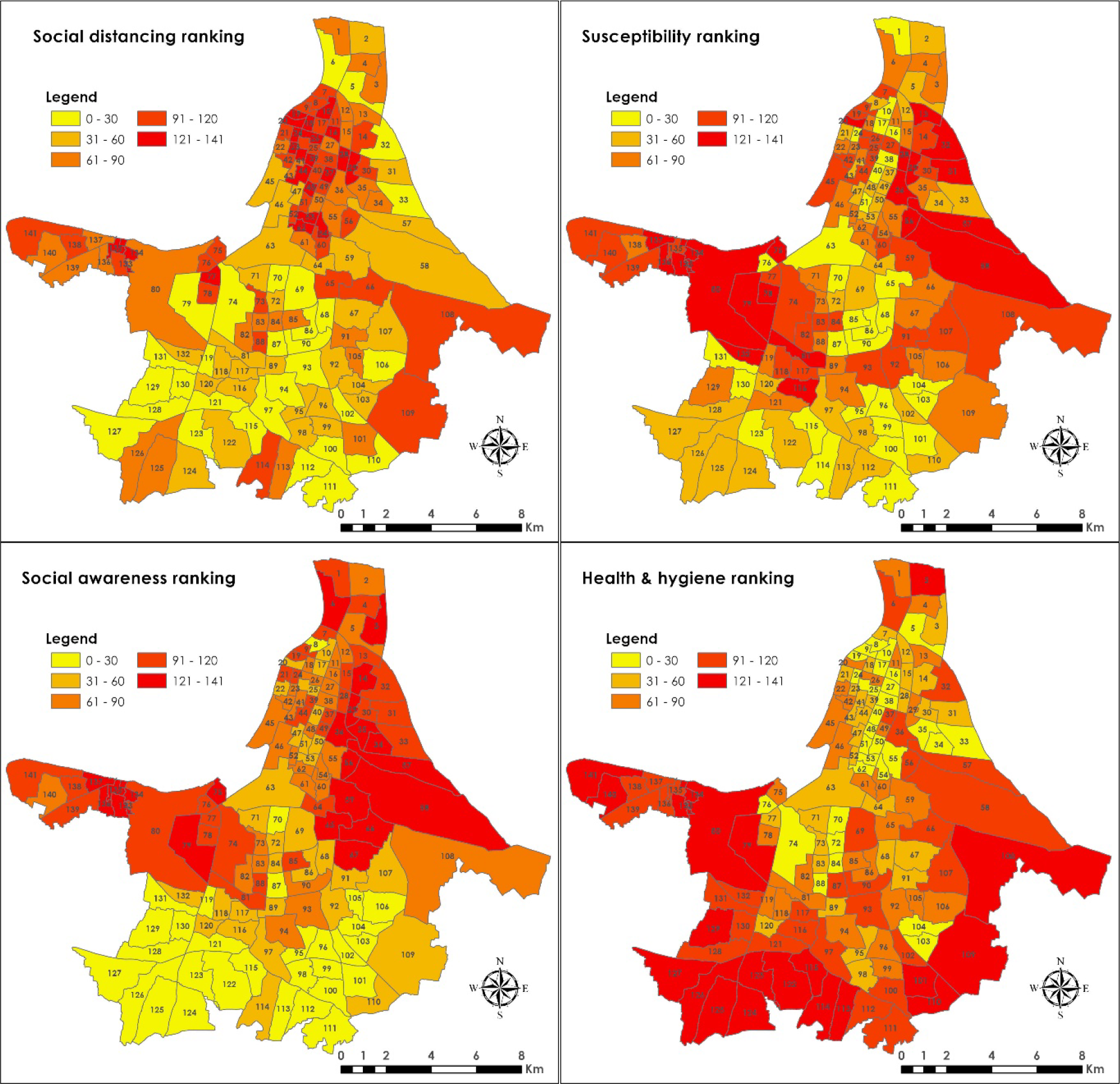
Ranking of KMC wards on the basis of objective driven four indices (lack of social distancing index, susceptibility index, lack of social awareness index, and poor of health & hygiene index) for the formulation of priority strategy(s) to prevent and control transmission of virus infections.

Susceptibility ranking indicate that the areas in east and west Kolkata have received largest ranking, which suggests that the majority of susceptible population are living in those localities. These are the areas where large fraction of wards population live below the poverty level and also having higher share of marginal workers (Fig. 5b and S1). These group of population are amongst the most vulnerable due to job losses and shutdown of economy. However, *r_R_* values indicate that the percentage of BPL population (*r_R_* = +0.95) has a more important role on overall rank weighted susceptibility index. Similarly, lack of social awareness index ranking show highest priority areas in the east and northeastern Kolkata and on the west of Kalighat, adjoining Alipore, Mominpore, Garden Reach and Metiabruz areas (Fig. 5c). Again, these localities house greater share of slum population and uneducated population. Likewise, *r_R_* values indicate that both the percentage of uneducated population (*r_R_* = +0.75) and slum population (*r_R_* = +0.94) have both played an important role on the ‘optimum rank’ of the lack of social awareness. Therefore, it would be important for the policy makers to make a plan for awareness campaign and community education about the risks associated with Covid-19 infections. Poor health and hygiene condition is one of the important factor for Covid-19 infections because the virus can attack easily the people with weak immune system. Keeping in mind that Covid-19 is a new virus and we have not developed any antibodies to fight against the virus, but people with weak immune system pose increased risk (https://www.cdc.gov/). According to health and hygiene index ranking, most of the highest priority areas are located in the east, south and west Kolkata (Fig. 5d). These are the areas where people have limited access to treated water, access to only community latrines, and the larger share of households with no wastewater drains (Fig. S1). However, the *r_R_* values indicate that the access to untreated water (*r_R_* = +0.72) and lack of wastewater drain (*r_R_* = +0.85) have both played an important role on the ‘optimum rank’ of the poor health & hygiene conditions.

### 3.5 Prevention and/or intervention strategies

We created a ranked based ‘predominance category map’ to find out what intervention category(s) should be followed in the hard hit wards (Fig. 6). The degree and/or strength of predominance category is highly dependent on how large is the rank difference and how high is the percent total of rank value. The data revealed that 4 out of 15 hard hit wards (ward no. 71, 72, 73 and 82) have all intervention criteria are almost equally important because all the objectives ranked equally largest. This suggests that multiple intervention objectives would be applicable to those wards to control the virus. While three criterion are matching in 4 wards (ward no. 7, 85, 86 and 94), two criterion are also matching in 4 wards (ward no. 13, 63, 74 and 76). Two criterion are matching in 2 wards (ward no. 68 and 70), however, ward no. 33 has a dominant intervention criterion of awareness. This suggests that other than ward no. 33, multiple factors are responsible for the presence of intense containment zones in these wards to stop virus transmission.

**Figure 6.**
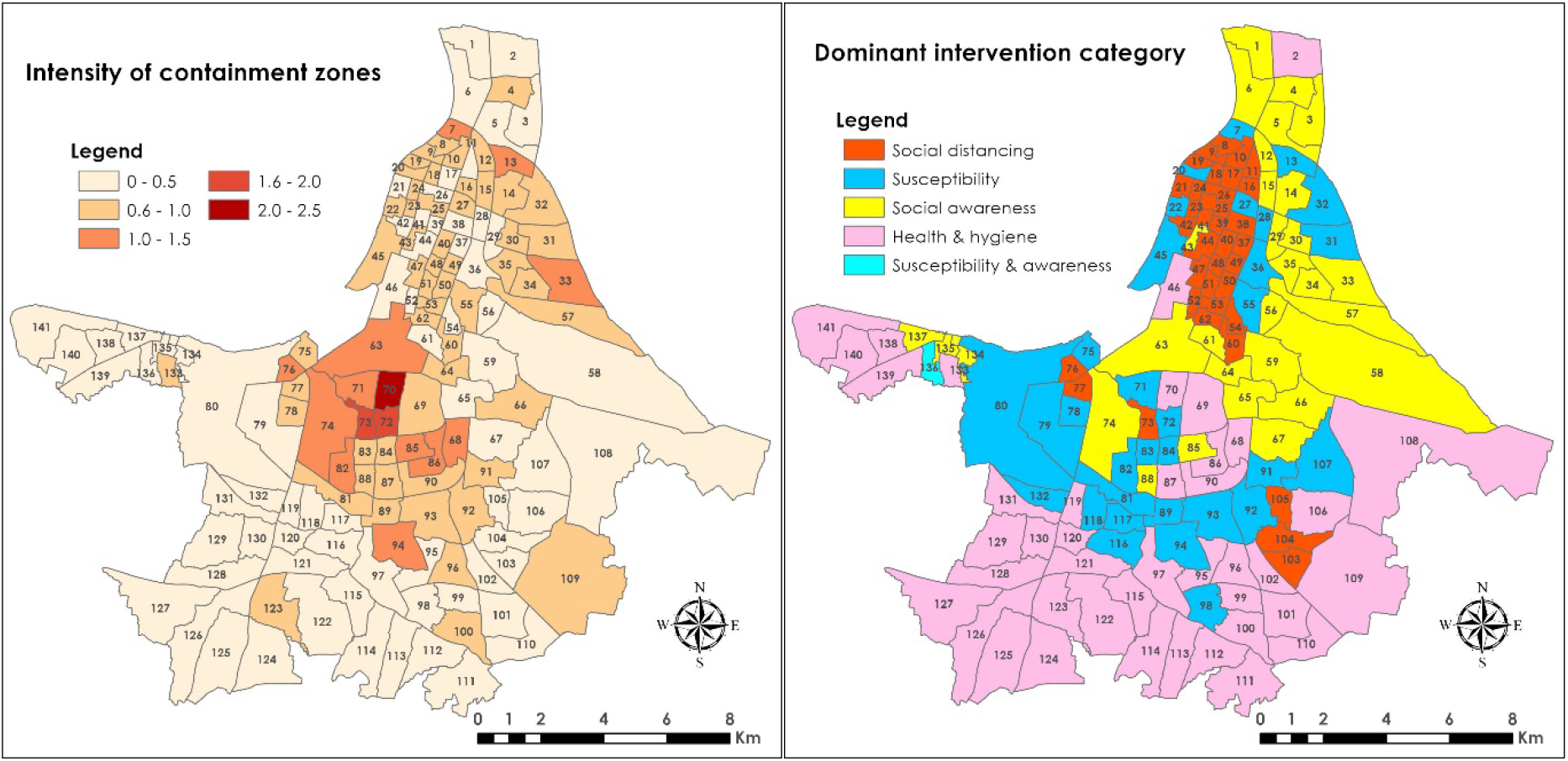
Comparison of the A) Distribution of hard hit areas within the KMC, and B) Dominant category of Covid-19 intervention criterion (the strength of predominance depends on the percent total and/or rank difference).

**Figure 7.**
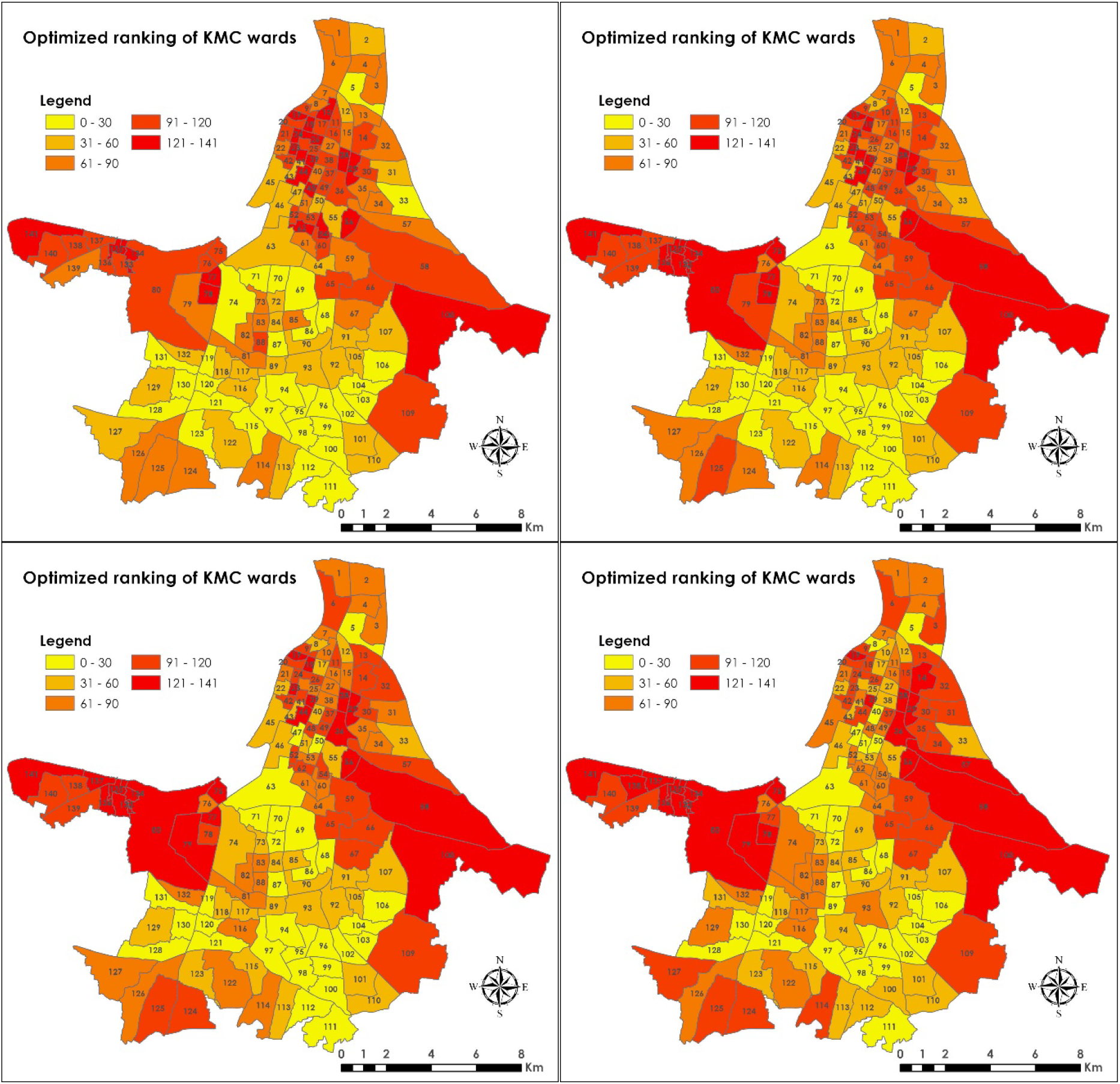
Future scenarios based on the optimized ranking of KMC wards: A) 0.7 weight to social distancing, and 0.1 weight equally to other three factors, B) 0.55 weight to social distancing, and 0.15 weight equally to other three factors, C) 0.4 weight to social distancing, and 0.2 weight equally to other three factors, and D) 0.25 weight to all four intervention criterion. Based on these optimized ranking targeted intervention on a priority basis to control Covid-19 infections/transmissions or in the case of future outbreak. Further work is needed to optimize the contributions of different weight factors.

Now it is important for the policy makers to adopt and chose priority planning program based on the degree of different risk factors. For example, if a ward ranked largest based on social distancing index, the planners could implement strict social distancing rule by restricting large gatherings and mass activities that could lead to higher human to human interactions and transmission of the disease. Assistance to those underprivileged population in the form of subsidy and/or financial packages during the pandemic would help ease their financial burden, because certain occupations pose greater risks of Covid-19 infections, especially those live in overcrowded places (https://www.ppic.org/blog/overcrowded-housing-and-covid-19-risk-among-essential-workers/). Temporary health care facilities could be made available in the highest priority areas to help deal with the Covid-19 infections. The Government of West Bengal have taken various initiatives to control the transmission of the virus by encouraging people to stay in home isolation and made thousands of containment zones that have contained for at least 14 days. In order to look after the vulnerable population, free ration to poor are provided till June, 2021 and food for those without ration card, migrant workers and homeless people. In addition to that special focus on shelter home to avoid transmission of disease due to lack of physical distancing and new shelter home for homeless sleeping in the street. Free mask and hand sanitizers have also provided by the officials, including teaching social distancing norms. However, even with such strong measures it is still difficult to contain the virus, perhaps due to lack of people’s awareness’s and reluctant to follow guidelines recommended by the administration. Because even a slight lapses in terms of taking precautions and following guidelines could be deadly. For example, allowing strangers and/or visiting friends would pose a greater risk to your health (https://www.texmed.org/TexasMedicineDetail.aspx?id=53977).

## 4. Implications for current and future scenarios

The analysis of broad-based containment zones listed for July and August revealed an interesting pattern (Fig. 3c). The majority of these containment zones are apartment complexes where people live in high rise buildings. There could be a possibility that the residents of the apartment complexes might not have maintained social distancing guidelines because of limited spaces available with apartment complexes. In addition to that, these apartment complexes need maintenance workers from outside and likewise the chances of transmission in under such circumstances should have increased significantly. According to a report, almost 50% of Covid-19 cases were reported from apartments during the first 25 days of July (TOI, 2020a).

We used Spearman’s rank correlation coefficient to ‘optimize the ranking’ of different wards based on the individual rankings of social distancing problem, awareness issues, susceptibility, and poor health & hygiene condition. A ‘weighted average’ approach was adopted to evaluate the impact of individual risk factors to the optimized rankings of wards in order of priority (high to low) and compare the results with current scenarios and its usefulness in the future. A ‘weighted average value’ for each risk factors was chosen based on the current understanding that the adherence to social distancing would significantly slow the spread of the virus so that healthcare system would not be strained. While, other factors such as susceptibility and poor health & hygiene conditions would have lesser impact in comparison to social distancing regulations. However, awareness to the Covid-19 risk could be as important as social distancing. Instead, we chose to emphasize social distancing because Kolkata is one of the most crowded metropolitan city in the world and therefore maintaining social distancing would a huge task for the administration and the local people. We therefore ranked all the wards based 0.25, 0.4, 0.55 and 0.7 weight given to social distancing and keeping the remaining 3 factors as equal weight, considering the total weighted value to be 1.

The results indicate that while considering 0.7 weight to social distancing and 0.1 weight each to awareness, susceptibility, and health & hygiene factors, the largest ranked wards are mostly located in the north and west of the city, while some wards in east of the city also ranked the largest. We found 9 out of 40 wards have more than 20 containment zones have ranked in the top 40 in optimized ranking. The number of wards remained the same even after reducing the weight to 0.55 for social distancing and increasing weight to 0.15 for all other factors. However, total number of high ranked wards have increased to 12 and 14, respectively, when social distancing contribution have decreased to 0.4 and 0.25, respectively. We then used other combination of weight such as giving 0.4 weight to awareness and 0.2 weight to the remaining and 0.4 weight to susceptibility and 0.2 weight to the remaining. The optimized ranking for awareness 0.4 weight suggests that 15 out of 40 highest ranked wards have more than 20 containment zones, while 16 out of 40 highest ranked wards have more than 20 containment zones. This suggests that social distancing is not essentially controlling the transmission of Covid-19 infections in different parts of Kolkata. However, a combination of other factors such as lack of awareness and susceptibility are likely contributing more to the current condition of virus transmission in Kolkata. Effectively three most important things is necessary follow, i.e., wearing masks, keeping a safe distance and maintaining hand hygiene that effectively minimizes the risks to Covid-19 transmission.

Health and social inequities can lead to increased risk of infection, such as discrimination, occupation, access to healthcare facilities, and choices in job situation. Lower socio-economic status together with poor health & hygiene condition may exacerbate Covid-19 infections. According to a study in peri-urban Tamil Nadu, India, the study population perceived no or low risk of contracting virus instead they feared for economic losses due to this pandemic (Kuang et al., 2020). A considerable proportion of this community did not increase their practice of preventive behaviors, such as social distancing and restrict gatherings. The similar scenario could a possibility in case of Kolkata Municipal Corporation. Therefore, raising awareness among public would benefits from widespread infections from coronavirus. In addition to that the government should be more proactive to address the needs of the population.

These ‘optimized ranked’ map can be used by the policy makers to implement various preventive measures and make resources available to control virus infections. We strongly believe that the policy makers and government agencies could follow our analysis protocol and make planning using better access to the data on Covid-19 infections in different KMC wards and follow informed judgement on selecting priority wards for interventions.

## 5. Conclusion

The analysis of Covid-19 risk factors based on the available data suggests various preventive and planning scenarios. The maps provided a detailed visualization of the potential issues that are important to be addressed and need to be discussed in detail to reduce the risk of contracting Covid-19 in the future. Additionally, identification of high risk and hard hit wards along with associated community characteristics enabled formulation of planning objectives for the future to minimize the impact of Covid-19 infections. We found that social distancing alone is not causing the transmission of virus in Kolkata, instead a combination of multiple risk factors playing a critical role in the increasing number of Covid-19 cases. Our results indicate that raising awareness among the population living in the high risk areas, e.g., increase use of masks, hand hygiene and keeping safe distance, would benefit the control of virus spread.

## Data Availability

We will make the data available on request.

